# Burden of insomnia on healthcare utilization in children with autism spectrum disorder

**DOI:** 10.1101/2022.12.14.22283459

**Authors:** Shirley Solomon, Gal Meiri, Analya Michaelovski, Yair Sadaka, Michal Ilan, Michal Faroy, Ilan Dinstein, Idan Menashe

## Abstract

Insomnia is frequently reported in children with autism spectrum disorder (ASD) and is associated with the severity of hyperactivity, irritability, sensory sensitivities, and related symptoms. The aim of this study was to examine the extent of healthcare utilization associated with insomnia in children with ASD. We conducted a retrospective, cross-sectional study of 541 children with ASD registered at the National Autism Database of Israel (NADI). Parents of all children completed the Children’s Sleep Habits Questionnaire (CSHQ) and insomnia was defined as a total CSHQ score ≥48. We compared sociodemographic characteristics, ASD diagnostic measures, chronic comorbidities, medication usage, hospitalizations, visits to the emergency room (ER), and visits to specialists between ASD children with and without insomnia. Finally, we assessed the independent association of insomnia with clinical characteristics and healthcare utilization using multivariate logistic regression models. Of the 541 children with ASD, 257 (47.5%) had insomnia. Children with insomnia compared to children without insomnia exhibited higher rates of comorbidities within the symptoms, signs, and ill-defined conditions (ICD-9[780–789]) category (aOR=1.70; 95%CI=1.04-2.77; p=0.033) and were prescribed more medications for chronic comorbidities (aOR=1.47; 95%CI=1.01-2.15; p=0.046). Finally, ASD children with insomnia were 1.75 and 2.82 times more likely to visit the ER and be hospitalized than their counterparts (aOR=1.75; 95%CI=1.17-2.62; p=0.007 and aOR=2.82; 95%CI=1.43-5.56; p=0.003, respectively). Our findings demonstrate that insomnia is associated with greater healthcare utilization among children with ASD. Thus, treating insomnia in children with ASD may have a broad clinical impact beyond specific improvements in sleep disturbances.

## Introduction

Adequate sleep is essential for normal brain development in children (Ednick et al., 2009) and reduces the risk of mental health disorders (Samanta et al., 2020), hypertension (Grandner et al., 2018), cardiovascular disease (Gottlieb, Somers, Punjabi, & Winkelman, 2017), obesity, and type-2 diabetes (Reutrakul & Van Cauter, 2018) into adulthood. In contrast, insomnia, which is defined as difficulty in initiating or maintaining sleep over a sustained period of time (American Psychiatric Association & Task Force, 2013), is associated with higher usage of medications, more frequent physician visits, and twice as many hospitalizations in the general population (Léger, Guilleminault, Bader, Lévy, & Paillard, 2002).

Insomnia, identified using parent questionnaires, is reported in 40-80% of children with autism spectrum disorder (ASD) (Richdale & Schreck, 2009) in contrast to approximately 25% in typically developing children (Owens & Maski, 2016). The types of sleep disturbances reported in children with ASD include, but are not limited to, difficulties in the initiation and maintenance of sleep, restless sleep, compromised sleep quality, frequent awakenings, and a total reduction in sleep time that typically begins during the first or second year of life (Dominick, Davis, Lainhart, Tager-Flusberg, & Folstein, 2007).

Several studies have suggested that sleep disturbances in children with ASD occur due to circadian rhythm abnormalities and, more specifically, disrupted secretion of melatonin (Miano et al., 2007). Alternatively, sleep disturbances in children with ASD may be caused by disrupted sleep homeostasis such that the pressure to sleep may be reduced relative to controls (Arazi et al., 2020). Melatonin supplementation is one of the most common treatments for insomnia in ASD (Schroder et al., 2021) with several studies reporting that both immediate release (IR) and prolonged release (PR) melatonin effectively facilitate sleep initiation. PR melatonin has been shown to increase total sleep duration by up to 62 minutes while IR melatonin increases total sleep time by 40.5 minutes (Dowling et al., 2005; Ferracioli-Oda, Qawasmi, & Bloch, 2013; P. Gringras et al., 2012; Paul Gringras, Nir, Breddy, Frydman-Marom, & Findling, 2017; Maras et al., 2018).

Sleep disturbances have been associated with the presentation of additional behavioral symptoms in children with ASD. For example, several studies reported that children with ASD and insomnia also manifest higher sensory sensitivities (Cortesi, Giannotti, Ivanenko, & Johnson, 2010; Manelis-Baram et al., 2021; Tzischinsky et al., 2018) and more severe aberrant behaviors including irritability, hyperactivity, inattention, and hostility (Hollway, Aman, & Butter, 2013; Mazurek & Sohl, 2016). Shorter sleep duration has also been associated with social communication impairment and increased restricted and repetitive behaviors (RRBs) (Veatch et al., 2017). This has led to the suggestion that insomnia may exacerbate the severity of core and secondary ASD symptoms and that treating insomnia may help reduce these symptoms (Lord, 2019). Indeed, one recent study has reported that treatment with PR melatonin improved total sleep time and consequently reduced hyperactivity in children with ASD (Schroder et al., 2019).

Given the high prevalence of insomnia in children with ASD, the goal of the current study was to determine the extent of healthcare utilization associated with insomnia in children with ASD. We utilized data from the National Autism Database of Israel (NADI) at the Azrieli National Center for Autism and Neurodevelopment Research to compare the prevalence of chronic comorbidities and the amount of health service utilization between ASD children with and without insomnia.

## Methods

### Participants

We conducted a retrospective, cross-sectional study of 541 children with ASD, between the ages of 1-11 years old, who were registered at the NADI (Dinstein et al., 2020; Meiri et al., 2017) between 2015 and 2021. Children were included in the current study if their parents completed the Children’s Sleep Habits Questionnaire (CSHQ) and if they were members of Clalit Health Services (CHS). CHS is the largest health maintenance organization (HMO) in Israel which insures 70% of the population in the south of Israel. We focused solely on members of CHS, because the children’s electronic patient records from this HMO were available to us through the Soroka University Medical Center’s (SUMC) medical database. The study was approved by the SUMC Helsinki committee.

### Evaluation of Insomnia

Sleep disturbances were evaluated using the CSHQ, a 33-item parent-rated questionnaire. The CSHQ yields scores in eight subscales relating to common types of sleep disturbances: bedtime resistance, sleep onset delay, sleep duration, anxiety around bedtime, parasomnias, night wakening, sleep-disordered breathing, and daytime sleepiness. All items are summed to create a final total score ranging between 33 and 99, with higher scores indicating greater severity. Insomnia was defined as a total sleep disturbance score ≥48, which is a conservative threshold for identifying children with severe sleep disturbances (Reynolds et al., 2019).

### Evaluation of core and secondary ASD symptoms

Core ASD symptom severity was evaluated in all children using the Autism Diagnostic Observation Schedule, 2^nd^ edition (ADOS-2) calibrated severity score (CSS). The ADOS-CSS, computed from ADOS-2 raw scores, allows comparison of ADOS-2 total scores across ages and modules (Gotham, Pickles, & Lord, 2009). We also compared DSM-5 levels of required support in the social communication (category A) and restricted, repetitive behaviors (RRB; category B) domains (American Psychiatric Association & Task Force, 2013). In addition, cognitive assessment scores from either the Bayley scales of infant and toddler development (Bayley, 2006) or the Wechsler Preschool and Primary Scale of Intelligence (WPPSI) (Wechsler, 1967) were available for most (n=445, 82%) of the children included in the final study sample.

### Evaluation of Health services utilization and medication use

Health records were obtained for all participating children from the CHS electronic patient record system. We extracted all chronic comorbidities, which were coded according to the International Classification of Diseases, Ninth Revision (ICD-9) format, and grouped them into broader disease categories (excluding complications of pregnancy, childbirth, and the puerperium [630-676] and conditions originating in the perinatal period [760-799]). Records of medication usage were obtained and grouped based on primary clinical use (Supplementary **Table S1**). In addition, hospitalizations, visits to the emergency room, and visits to specialists during a time period corresponding to one year before and after completion of the CSHQ were also gathered from the electronic records.

### Statistical Analyses

Standard univariate tests were used to examine differences in various demographic and clinical characteristics between children with ASD, with and without insomnia. Specifically, chiΔsquare or Fisher-exact tests were used to assess for differences in categorical variables, and Mann–Whitney UΔtest for continuous variables. Finally, multivariable logistic regression models were used to assess the independent association of each of these variables with insomnia, adjusted for basic sociodemographic and clinical covariates (e.g. age, sex, ethnicity, and DSM-V B required level of support). A p-value of <0.05 was considered statistically significant. The statistical analyses were performed using R studio, version 1.4.1717 (R Foundation for Statistical Computing version).

## Results

Of the 1,108 children with ASD in the NADI as of August 2021, 541 children (48.9%) fulfilled the study inclusion criteria. Children included in the sample did not differ significantly from those who were not with respect to age at diagnosis, sex, ethnic origin, cognitive scores, and ADOS-2 calibrated severity scores (**Table 1**). Children in the study, however, required more support compared to children who were not included in the study sample, as estimated by the diagnosing physician according to the DSM-5 levels of required support. This difference may reflect a tendency of parents of children who require more support, to complete the CSHQ questionnaire.

**Table 1.**
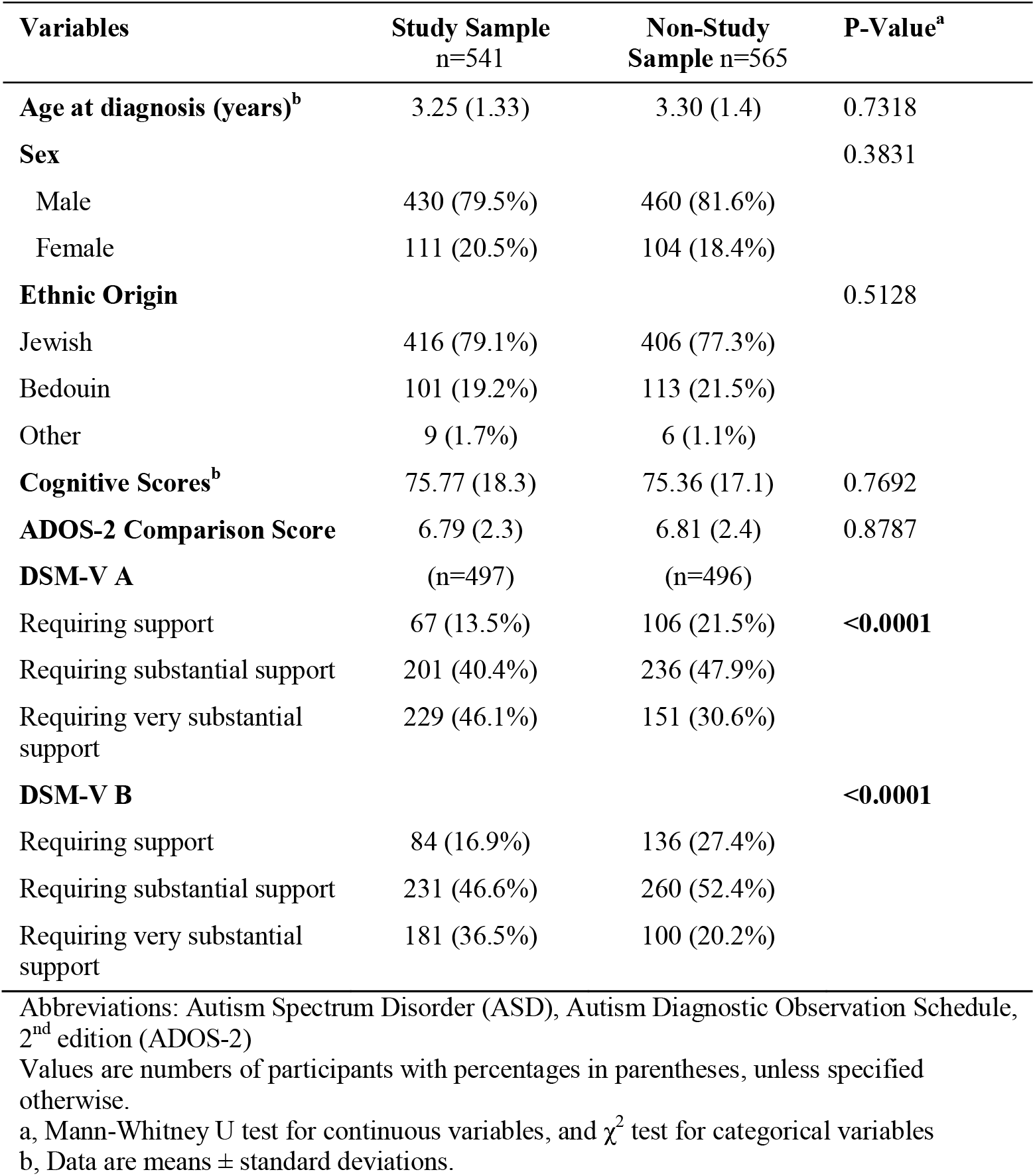
Comparison of characteristics across children with ASD registered in the National Autism Database of Israel who met inclusion criteria for the current study versus those who did not.

| **Variables** | **Study Sample**<br>**n=541** | **Non-Study**<br>**Sample n=565** | **P-Value<sup>a</sup>** |
| --- | --- | --- | --- |
| **Age at diagnosis (years)<sup>b</sup>** | 3.25 (1.33) | 3.30 (1.4) | 0.7318 |
| **Sex** |  |  | 0.3831 |
| Male | 430 (79.5%) | 460 (81.6%) |  |
| Female | 111 (20.5%) | 104 (18.4%) |  |
| **Ethnic Origin** |  |  | 0.5128 |
| Jewish | 416 (79.1%) | 406 (77.3%) |  |
| Bedouin | 101 (19.2%) | 113 (21.5%) |  |
| Other | 9 (1.7%) | 6 (1.1%) |  |
| **Cognitive Scores<sup>b</sup>** | 75.77 (18.3) | 75.36 (17.1) | 0.7692 |
| **ADOS-2 Comparison Score** | 6.79 (2.3) | 6.81 (2.4) | 0.8787 |
| **DSM-V A** | (n=497) | (n=496) |  |
| Requiring support | 67 (13.5%) | 106 (21.5%) | **<0.0001** |
| Requiring substantial support | 201 (40.4%) | 236 (47.9%) |  |
| Requiring very substantial support | 229 (46.1%) | 151 (30.6%) |  |
| **DSM-V B** |  |  | **<0.0001** |
| Requiring support | 84 (16.9%) | 136 (27.4%) |  |
| Requiring substantial support | 231 (46.6%) | 260 (52.4%) |  |
| Requiring very substantial support | 181 (36.5%) | 100 (20.2%) |  |

Abbreviations: Autism Spectrum Disorder (ASD), Autism Diagnostic Observation Schedule, 2<sup>nd</sup> edition (ADOS-2)
Values are numbers of participants with percentages in parentheses, unless specified otherwise.
a, Mann-Whitney U test for continuous variables, and $\chi^2$ test for categorical variables
b, Data are means ± standard deviations.

Overall, participating children had a broad distribution of CSHQ scores ranging from 33 to 81, with 257 (47.5%) of them exhibiting CSHQ scores ≥ 48 indicative of insomnia (**Figure 1**). There were no significant differences in sex ratio, cognitive scores, or ADOS-2 calibrated severity scores across children with and without insomnia (**Table 2**). However, there were more children of Bedouin origin in the insomnia group (24.6% vs. 14.2%; p=0.0101) and the mean age of the children in the insomnia group was slightly higher (4.34 vs. 4.03 years; p=0.0394). In addition, children with insomnia required more support according to the DSM-V criteria with significant differences observed in the B criteria describing difficulties in RRB symptoms (p=0.0278).

**Table 2.** Comparison of characteristics across children with and without insomnia

| **Sociodemographic Variables** | **Non-Insomnia**<br>**(CHSQ<48)**<br>**n=284** | **Insomnia**<br>**(CHSQ≥48)**<br>**n=257** | **P-Value<sup>a</sup>** |
| --- | --- | --- | --- |
| **Age (years)<sup>b</sup>** | 4.03 (1.8) | 4.34 (1.8) | **0.0394** |
| **Sex** |  |  | 0.8763 |
| Male | 225 (79.2%) | 205 (79.8%) |  |
| Female | 59 (20.8%) | 52 (20.2%) |  |
| **Ethnic Origin** |  |  | **0.0101** |
| Jewish | 230 (83.9%) | 186 (73.8%) |  |
| Bedouin | 39 (14.2%) | 62 (24.6%) |  |
| Other | 5 (1.8%) | 4 (1.6%) |  |
| **Cognitive Scores<sup>b</sup>** | 76.46 (17.2) | 74.92 (19.6) | 0.5275 |
| **ADOS-2 CSS** | 6.67 (2.3) | 6.94 (2.3) | 0.1906 |
| **DSM-V A** |  |  |  |
| Mild | 36 (14.0%) | 31 (13.0%) | 0.1955 |
| Moderate | 112 (43.4%) | 89 (37.2%) |  |
| Severe | 110 (42.6%) | 119 (49.8%) |  |
| **DSM-V B** |  |  | **0.0278** |
| Mild | 47 (18.3%) | 37 (15.5%) |  |
| Moderate | 130 (50.6%) | 101 (42.3%) |  |
| Severe | 80 (31.1%) | 101 (42.3%) |  |

Abbreviations: Autism Spectrum Disorder (ASD), Autism Diagnostic Observation Schedule, 2<sup>nd</sup> edition (ADOS-2), Children's Sleep Habits Questionnaire (CSHQ), Values are numbers of participants with percentages in parentheses, unless specified otherwise.
a, Mann-Whitney U test for continuous variables, and $\chi^2$ test for categorical variables
b, Data are means ± standard deviations.

**Figure 1.**
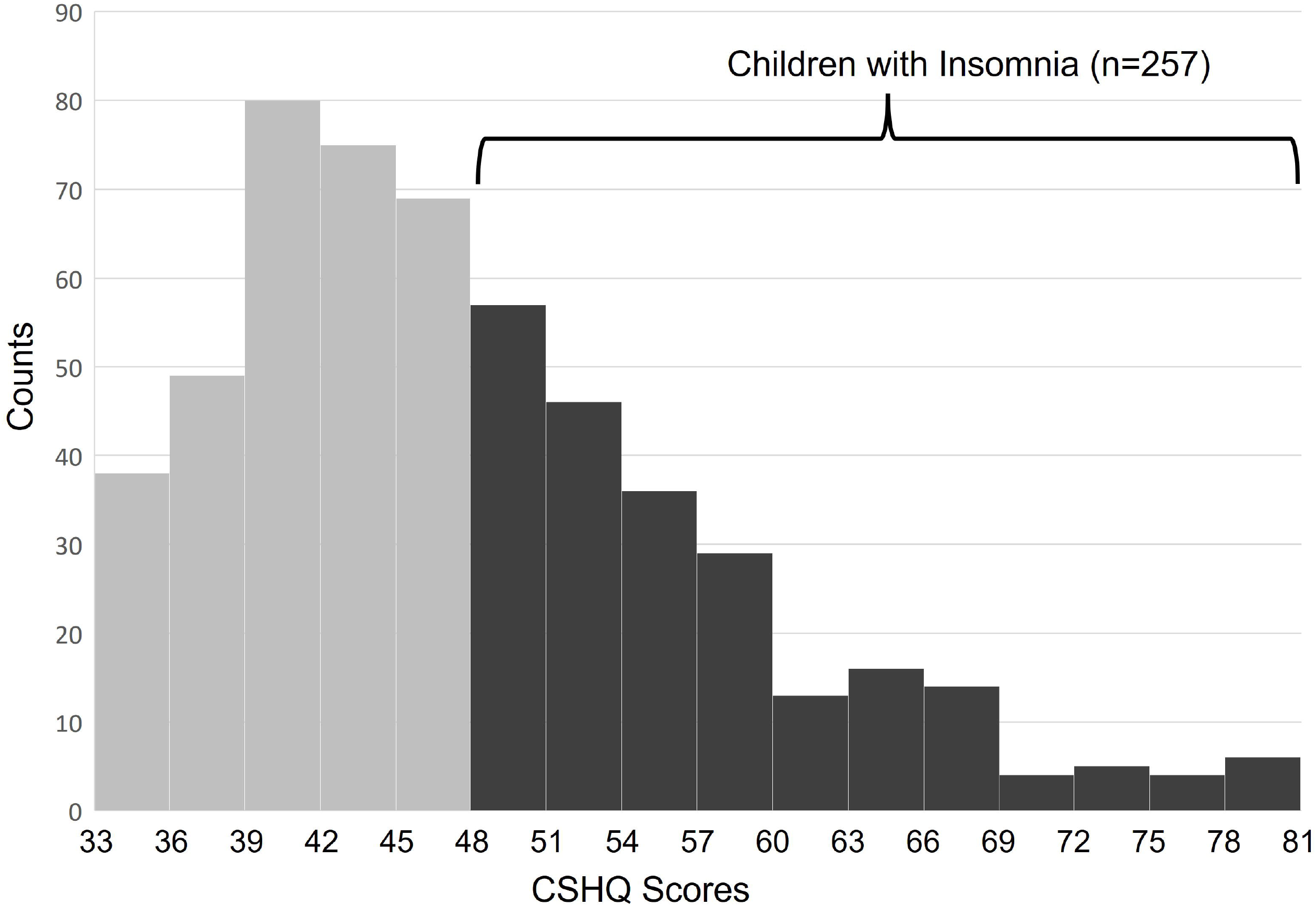
Distribution of CSHQ scores in the study sample. A histogram of the frequency of CSHQ scores (X-axis) of children with ASD in the study sample. Overall, 257 children had CSHQ scores ≥48 and were defined as having insomnia. Abbreviations: Children’s Sleep Habits Questionnaire (CSHQ)

### Chronic comorbidities

ASD Children with insomnia had significantly more comorbidities from the symptoms, signs, and ill-defined conditions classification than ASD children without insomnia (**Table 3**; 21.0% vs. 13.7%, P = 0.0334). There were no significant differences in other examined categories of comorbidities. There was no significant difference in number of ASD children who had comorbidities (49.8% vs. 45.8%; p = 0.3947), although those with insomnia tended to have a larger number of coexisting comorbidities than their counterparts. Notably, 19.1% of children with insomnia had three or more comorbidities, compared to only 12.7% of children without insomnia (p = 0.0414).

**Table 3.** Comparison of chronic comorbidities between ASD children with and without insomnia

|  |  | **Non-Insomnia**<br>**(CHSQ<48)**<br>**n=284** | **Insomnia**<br>**(CHSQ≥48)**<br>**n=257** | **P-Value<sup>\*</sup>** |
| --- | --- | --- | --- | --- |
| **Any Chronic Comorbidity** |  | 130 (45.8%) | 128 (49.8%) | 0.3947 |
| **Number of Chronic Comorbidities** | **1** | 59 (20.8%) | 50 (19.5%) | 0.7024 |
|  | **2** | 35 (12.3%) | 29 (11.3%) | 0.7084 |
|  | **≥ 3** | 36 (12.7%) | 49 (19.1%) | **0.0414** |
| Infectious and Parasitic |  | 3 (1.1%) | 2 (0.8%) | 1 |
| Neoplasms |  | 0 (0.0%) | 1 (0.4%) | 0.4750 |
| Endocrine, Nutritional, Metabolic, and Immunity |  | 14 (4.9%) | 15 (5.8%) | 0.7821 |
| Blood and Blood-Forming Organs |  | 16 (5.6%) | 13 (5.1%) | 0.9159 |
| Mental Disorders |  | 15 (5.3%) | 22 (8.6%) | 0.1808 |
| Nervous System and Sense Organs |  | 31 (10.9%) | 35 (13.6%) | 0.4078 |
| Circulatory System |  | 1 (0.4%) | 1 (0.4%) | 1 |
| Respiratory System |  | 29 (10.2%) | 33 (12.8%) | 0.4102 |
| Digestive System |  | 5 (1.8%) | 8 (3.1%) | 0.4565 |
| Genitourinary System |  | 2 (0.7%) | 5 (1.9%) | 0.2653 |
| Skin and Subcutaneous Tissue |  | 9 (3.2%) | 16 (6.2%) | 0.1373 |
| Musculoskeletal System and Connective Tissue |  | 4 (1.4%) | 5 (1.9%) | 0.7421 |
| Congenital Anomalies |  | 15 (5.3%) | 22 (8.6%) | 0.1808 |
| Symptoms, Signs, And Ill-Defined Conditions |  | 39 (13.7%) | 54 (21.0%) | **0.0334** |
| Injury and Poisoning |  | 24 (8.5%) | 25 (9.7%) | 0.7138 |
| *Developmental Delay* |  | 22 (7.7%) | 21 (8.1%) | 0.9815 |

Abbreviations: Autism Spectrum Disorder (ASD), Children's Sleep Habits Questionnaire (CHSQ)
Values are numbers of participants with percentages in parentheses.
<sup>\*</sup> $\chi^2$ test or Fisher exact tests were conducted to determine the association between chronic comorbidities and CHSQ score.

### Medications use

Use of medication for the management of chronic diseases (as listed in Supplementary **Table S1**) for both groups is presented in **Table 4**. Overall, children with insomnia were more likely to be prescribed medications than children without insomnia (45.5% vs. 32.7%, P = 0.0031). This difference was partially due to prescription of medication for insomnia (e.g., Melatonin and Promethazine) that was almost twice more frequent in the insomnia group (15.2% vs. 8.1%, P=0.0145). Nevertheless, the amount of prescribed medications for children with ASD and insomnia was still significantly higher after excluding medications that treat insomnia (37.0% vs. 28.2%, P = 0.0364), demonstrating that they use more medications that are unrelated to sleep. In particular, prescription for medications in the treatment of mental or mood conditions were significantly more frequent in the insomnia group (9.7% vs. 4.9%, P = 0.0468).

**Table 4.** Comparison of medication usage between children with and without insomnia

| **Prescribed Medications<sup>a</sup>** | **Non-Insomnia**<br>**(CHSQ<48)**<br>**n=284** | **Insomnia**<br>**(CHSQ≥48)**<br>**n=257** | **P-Value** |
| --- | --- | --- | --- |
| **Any Medications** | 93 (32.7%) | 117 (45.5%) | **0.0031<sup>b</sup>** |
| **Insomnia** | 23 (8.1%) | 39 (15.2%) | **0.0145<sup>b</sup>** |
| **Medications, excluding those**<br>**for insomnia** | 80 (28.2%) | 95 (37.0%) | **0.0364<sup>b</sup>** |
| **Asthma/Pulmonary Disorders** | 30 (10.6%) | 30 (11.7%) | 0.7845<sup>b</sup> |
| **ADHD** | 30 (10.6%) | 26 (10.1%) | 0.9769<sup>b</sup> |
| **Mental and Mood Conditions** | 14 (4.9%) | 25 (9.7%) | **0.0468<sup>b</sup>** |
| **Nutrient Deficiencies** | 14 (4.9%) | 20 (7.8%) | 0.2349<sup>b</sup> |
| **Food supplements** | 9 (3.2%) | 16 (6.2%) | 0.1373<sup>b</sup> |
| **Seizure Disorders** | 6 (2.1%) | 8 (3.1%) | 0.6451<sup>b</sup> |
| **Skin Conditions** | 2 (0.7%) | 7 (2.7%) | 0.0927<sup>c</sup> |
| **Allergic Disorders** | 2 (0.7%) | 4 (1.6%) | 0.4303<sup>c</sup> |
| **Endocrine and Metabolic**<br>**Diseases** | 2 (0.7%) | 4 (1.6%) | 0.4303<sup>c</sup> |
| **Gastrointestinal Disorders** | 2 (0.7%) | 2 (0.8%) | 1<sup>c</sup> |
| **Pre-Surgical Medications** | 4 (1.4%) | 2 (0.8%) | 0.6881<sup>c</sup> |
| **Infections** | 0 (0%) | 3 (1.2%) | 0.1065<sup>c</sup> |
| **Urologic Disorders** | 3 (1.1%) | 0 (0%) | 0.2505<sup>c</sup> |
| **Autoimmune diseases** | 0 (0%) | 2 (0.8%) | 0.2252<sup>c</sup> |
| **Cardiovascular Diseases** | 2 (0.7%) | 0 (0%) | 0.5003<sup>c</sup> |
| **Ophthalmologic Conditions** | 1 (0.4%) | 1 (0.4%) | 1<sup>c</sup> |
| **Rheumatic diseases** | 1 (0.4%) | 1 (0.4%) | 1<sup>c</sup> |
| **Otorhinolaryngology**<br>**Conditions** | 1 (0.4%) | 0 (0%) | 1<sup>c</sup> |
| **Pain Medications** | 0 (0%) | 1 (0.4%) | 0.4750<sup>c</sup> |
| **Various Main Indications** | 1 (0.4%) | 0 (0%) | 1<sup>c</sup> |

Abbreviations: Autism Spectrum Disorder (ASD), Children's Sleep Habits Questionnaire (CSHQ), Attention Deficit Hyperactivity Disorder (ADHD)
Values are numbers of participants with percentages in parentheses.
<sup>a</sup> Medications were grouped based on primary clinical use (Supplementary Table S1).
<sup>b</sup> Pearson Chi-Square test. <sup>c</sup> Fisher-Exact test.

### Health services utilization

Children with ASD and insomnia utilized more health services than children without insomnia (**Figure 2**) including 50% more visits to the emergency room (mean[SD] = 0.63[1.19] vs. 0.42[1.01]; p= 0.0153) and a 2.7 times higher rate of hospitalization (mean[SD] = 0.19[0.60] vs. 0.07[1.30]; p= 0.0042). Consequently, children with insomnia were hospitalized for twice the number of days compared to children without insomnia (mean[SD] = 0.32[1.08] vs. 0.16[1.06] days per child respectively; p= 0.004). These findings suggest that the cost of emergency room visits and hospitalization is significantly higher for children with insomnia. No significant differences were found in the total number of outpatient visits, including visits to primary care physicians (mean[SD] = 16.7 [14.7] vs. 16.9 [13.2] visits per child respectively; p= 0.7492) and specialists (mean[SD] = 0.95[1.81] vs. 0.80[1.51] visits per child respectively; p= 0.5206).

**Figure 2.**
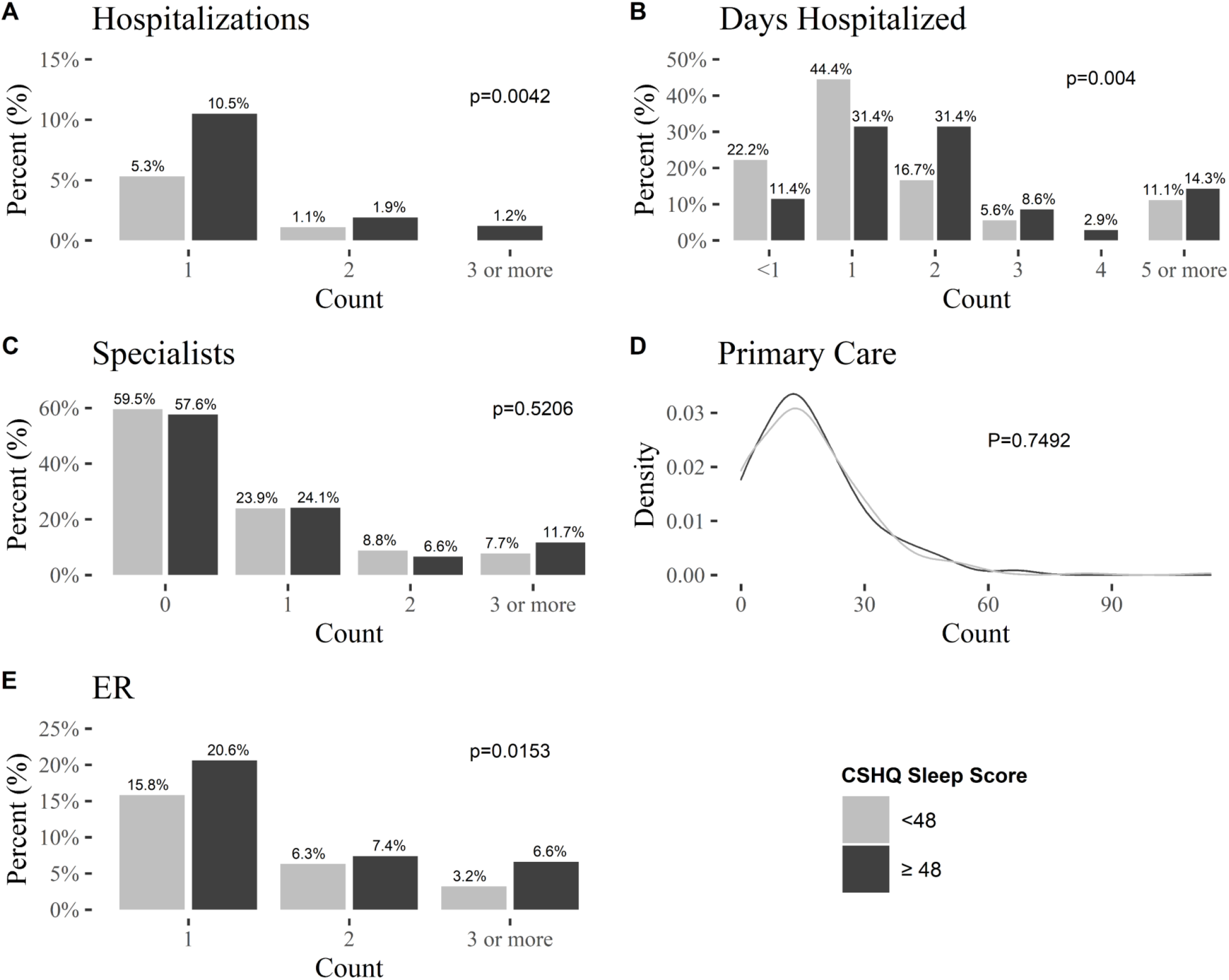
Percentage of health services utilization during a period of one year before and after completion of the CSHQ. **A**. Number of hospitalizations. **B**. Days hospitalized. **C**. Number of visits to a specialist. **D**. Number of visits to a primary care physician. **E**. Number of visits to the emergency room. Note that percentages presented in panels A, C and E sum to 13.6%, 42.4% and 34.6% of ASD children with insomnia and 6.4%, 58.5% and 25.3% of children without insomnia who were hospitalized, visited specialists, and who visited the emergency room, respectively. P-values from Mann-Whitney U tests are for the differences between children with and without insomnia. Abbreviations: Children’s Sleep Habits Questionnaire (CSHQ), Emergency room (ER),

### Factors associated with insomnia

Finally, we used multivariable logistic regression models, to quantify the independent association of factors associated with insomnia in our sample while controlling for potential confounders (**Table 5**). Insomnia in children with ASD was significantly associated with a 1.70 higher likelihood of having a comorbidity within the symptoms, signs, and ill-defined conditions classification (aOR = 1.70; 95% CI = 1.04, 2.77; p=0.033) and a 1.47 higher likelihood of medication use (aOR = 1.47; 95% CI = 1.01, 2.15; p=0.046). In addition, insomnia in children with ASD was also significantly associated with visits to the emergency room (≥1 visits during the study period) (aOR = 1.75; 95% CI = 1.17, 2.62; p=0.007), and hospitalizations (≥1 hospitalizations during the study period) (aOR = 2.82; 95% CI = 1.43, 5.56; p=0.003). No other health services factors were associated with insomnia in children with ASD after accounting for confounders.

**Table 5.** Factors associated with insomnia in children with ASD.

|  | **Crude OR**<br>(95% CI) | **Adjusted OR<sup>a</sup>**<br>(95%CI) |
| --- | --- | --- |
| **Chronic Comorbidities** | 1.18 (0.84, 1.65) | 1.04 (0.72, 1.51) |
| Symptoms, Signs, And Ill-Defined Conditions | 1.67 (1.06, 2.63)<sup>*</sup> | 1.70 (1.04, 2.77)<sup>*</sup> |
| Multiple comorbidities (3 or more) | 1.62 (1.02, 2.59)<sup>*</sup> | 1.58 (0.95, 2.63) |
| **Medication Use** | 1.72 (1.21, 2.43)<sup>*</sup> | 1.47 (1.01, 2.15)<sup>*</sup> |
| Medications for insomnia (Melatonin and Promethazine) | 2.03 (1.18, 3.50)<sup>*</sup> | 1.72 (0.96, 3.08) |
| Medications, excluding those for insomnia | 1.50 (1.04, 2.15)<sup>*</sup> | 1.28 (0.86, 1.90) |
| Mental and Mood Conditions | 2.08 (1.06, 4.09)<sup>*</sup> | 1.62 (0.75, 3.51) |
| **Emergency Room Visits (Yes/No)** | 1.56 (1.08, 2.26)<sup>*</sup> | 1.75 (1.17, 2.62)<sup>*</sup> |
| **Primary Care Visits (Yes/No)** | 1.00 (0.99, 1.01) | 1.00 (0.99, 1.02) |
| **Specialist Visits (Yes/No)** | 1.08 (0.77, 1.52) | 1.21 (0.84, 1.76) |
| **Hospitalizations (Yes/No)** | 2.33 (1.28, 4.23)<sup>*</sup> | 2.82 (1.43, 5.56)<sup>*</sup> |

Abbreviations: Autism Spectrum Disorder (ASD), Children's Sleep Habits Questionnaire (CSHQ), Odds Ratio (OR)
<sup>a</sup> Adjusted for sociodemographic variables (sex, ethnicity, age, and DSM-V category B).
<sup>\*</sup> P-value less than 0.05

## Discussion

This study examined the health burden associated with insomnia among children with ASD. The findings show that insomnia is present in nearly 50% of children with ASD and that these children have a higher number of comorbidities, use more medications for the management of chronic diseases, and are more likely to visit the emergency room and be hospitalized compared to children with ASD without insomnia. Altogether, these results suggest that insomnia is associated with a greater clinical and economic burden among children with ASD.

These findings from children with ASD are in line with previous studies that have reported positive associations between insomnia and a diverse range of comorbidities within the general adult population. These include associations between insomnia and anxiety and depression (Sørensen, Jensen, Rathleff, & Holden, 2019), gastrointestinal disorders (Balikji et al., 2018), psychopathy symptoms (Akram et al., 2018), and even a 45% increased risk of mortality from cardiovascular disease (Sofi et al., 2014). Some studies have proposed possible mechanistic explanations for the increased presence of comorbid disorders in individuals with insomnia including circadian rhythm misalignment (Nobre, Rocha, Morin, & Meira e Cruz, 2021), a low-grade inflammatory state (Friedman, 2011), elevation of cortisol (Vargas et al., 2018), and metabolic or endocrine changes (Copinschi, 2005). Some of these mechanisms were also suggested to be involved in ASD etiology (Abdul et al., 2022; Geoffray, Nicolas, Speranza, & Georgieff, 2016), thus highlighting potential mechanistic links between ASD and insomnia.

The association of insomnia with greater use of health services in this study is also consistent with previous reports of increased utilization of health services in adults with insomnia in the general population including more frequent emergency room visits and hospitalizations, but no significant increases in the number of physician visits (Léger et al., 2002; Skaer & Sclar, 2010). Moreover, in our study insomnia remained an independent predictor of healthcare utilization in ASD children even after controlling for the number of comorbidities in these children.

Finally, individuals with insomnia in the general population were also reported to consume more medications than good sleepers, especially, sleep medications, sedatives, anti-depressants, pain relief medications, asthma medications, and cardiac medications (Daley et al., 2009; Sivertsen, Krokstad, Mykletun, & Øverland, 2009). These results are also consistent with this study’s findings of higher overall medication use among ASD children with insomnia. Notably, in the current study, medications use for mental and mood conditions were also associated with insomnia in the unadjusted models. This finding is expected given that ASD children with insomnia display more aberrant behaviors (Hollway et al., 2013; Mazurek & Sohl, 2016), and antipsychotics, including risperidone and aripiprazole (the two most common medications prescribed in this study sample under the mental and mood conditions classification), are prescribed to reduce disruptive behaviors, particularly irritability and aggression in children with ASD (Ching & Pringsheim, 2012; Loy, Merry, Hetrick, & Stasiak, 2017). The higher rate of insomnia among children who are prescribed drugs for mental and mood conditions could be due to the known side effects of these drugs in causing sleep problems (Miller & McCall, 2022), however, the association of these drugs in our study were mild and not statistically significant (p = 0.220).

The results reported in this study should be interpreted in the context of the following limitations. First, information on the child’s sleep behavior was gathered via parental reports using the CSHQ. This method may be biased as it relies on parent impressions and their subjective ability to accurately recall their child’s sleep habits. Alternative methods such as daily sleep diaries or direct measures such as actigraphy and polysomnography may offer less biased estimates of sleep disturbances, however, these measures are often difficult to acquire from children with ASD due to sensory sensitivities and lack of cooperation (Goldman et al., 2009). Second, this study used a retrospective, cross-sectional design and thus it was not possible to determine causality or directionality of the association between the emergence of insomnia and utilization of health services. Lastly, data was obtained solely from the electronic records, which included only clinical data, and no information was obtained about use of unprescribed or over-the-counter medications, including melatonin.

## Conclusions

Our findings demonstrate that insomnia is significantly associated with greater healthcare utilization among children with ASD. First-line recommendations for insomnia treatment include a wide-range of behavioral interventions that can reduce sleep difficulties and improve sleep quality for some children with insomnia (Guénolé et al., 2011; Rossignol & Frye, 2011). In children with ASD, insomnia can be treated with behavioral interventions and then supplemented with pharmacological treatment (Schroder et al., 2021). Thus, future prospective follow-up studies could examine whether successful treatment of insomnia reduces health services utilization and comorbid disorders in ASD children. This would be critical for revealing whether insomnia treatments have a broad clinical impact beyond the reduction of sleep disturbances.

## Data Availability

All data produced in the present study are available upon reasonable request to the authors

## Acknowledgements

We thank the families who participated in this research.

## References

Abdul, F., Sreenivas, N., Kommu, J. V. S., Banerjee, M., Berk, M., Maes, M., … Debnath, M. (2022). Disruption of circadian rhythm and risk of autism spectrum disorder: role of immune-inflammatory, oxidative stress, metabolic and neurotransmitter pathways. Reviews in the Neurosciences, 33(1), 93–109. https://doi.org/10.1515/revneuro-2021-0022

Akram, U., Allen, S., McCarty, K., Gardani, M., Tan, A., Villarreal, D., … Akram, A. (2018). The relationship between insomnia symptoms and the dark triad personality traits. Personality and Individual Differences, 131, 212–215. https://doi.org/10.1016/j.paid.2018.05.001

American Psychiatric Association, & Task Force, D.-5. (2013). Diagnostic and Statistical Manual of Mental Disorders (DSM-5®). American Psychiatric Publishing. https://doi.org/https://doi.org/10.1176/appi.books.9780890425596

Arazi, A., Meiri, G., Danan, D., Michaelovski, A., Flusser, H., Menashe, I., … Dinstein, I. (2020). Reduced sleep pressure in young children with autism. Sleep, 43(6). https://doi.org/10.1093/sleep/zsz309

Balikji, S., Mackus, M., Brookhuis, K., Garssen, J., Kraneveld, A., Roth, T., & Verster, J. (2018). The Association of Insomnia, Perceived Immune Functioning, and Irritable Bowel Syndrome Complaints. Journal of Clinical Medicine, 7(9), 238. https://doi.org/10.3390/jcm7090238

Bayley, N. (2006). Bayley-III: Bayley Scales of infant and toddler development (3rd ed.). San Antonio, TX: Pearson.

Ching, H., & Pringsheim, T. (2012). Aripiprazole for autism spectrum disorders (ASD). In T. Pringsheim (Ed.), Cochrane Database of Systematic Reviews. Chichester, UK: John Wiley & Sons, Ltd. https://doi.org/10.1002/14651858.CD009043.pub2

Copinschi, G. (2005). Metabolic and endocrine effects of sleep deprivation. Essential Psychopharmacology, 6(6).

Cortesi, F., Giannotti, F., Ivanenko, A., & Johnson, K. (2010). Sleep in children with autistic spectrum disorder. Sleep Medicine, 11(7), 659–664. https://doi.org/10.1016/j.sleep.2010.01.010

Daley, M., Morin, C. M., LeBlanc, M., Grégoire, J. P., Savard, J., & Baillargeon, L. (2009). Insomnia and its relationship to health-care utilization, work absenteeism, productivity and accidents. Sleep Medicine, 10(4), 427–438. https://doi.org/10.1016/j.sleep.2008.04.005

Dinstein, I., Arazi, A., Golan, H. M., Koller, J., Elliott, E., Gozes, I., … Meiri, G. (2020, September). The National Autism Database of Israel: a Resource for Studying Autism Risk Factors, Biomarkers, Outcome Measures, and Treatment Efficacy. Journal of Molecular Neuroscience, Vol. 70, pp. 1303–1312. Humana Press Inc. https://doi.org/10.1007/s12031-020-01671-z

Dominick, K. C., Davis, N. O., Lainhart, J., Tager-Flusberg, H., & Folstein, S. (2007). Atypical behaviors in children with autism and children with a history of language impairment. Research in Developmental Disabilities, 28(2), 145–162. https://doi.org/10.1016/j.ridd.2006.02.003

Dowling, G. A., Mastick, J., Colling, E., Carter, J. H., Singer, C. M., & Aminoff, M. J. (2005). Melatonin for sleep disturbances in Parkinson’s disease. Sleep Medicine, 6(5), 459–466. https://doi.org/10.1016/j.sleep.2005.04.004

Ednick, M., Cohen, A. P., McPhail, G. L., Beebe, D., Simakajornboon, N., & Amin, R. S. (2009). A Review of the Effects of Sleep During the First Year of Life on Cognitive, Psychomotor, and Temperament Development. Sleep, 32(11), 1449–1458. https://doi.org/10.1093/sleep/32.11.1449

Ferracioli-Oda, E., Qawasmi, A., & Bloch, M. H. (2013). Meta-Analysis: Melatonin for the Treatment of Primary Sleep Disorders. PLoS ONE, 8(5), e63773. https://doi.org/10.1371/journal.pone.0063773

Friedman, E. M. (2011). Sleep quality, social well-being, gender, and inflammation: An integrative analysis in a national sample. Annals of the New York Academy of Sciences, 1231(1), 23–34. https://doi.org/10.1111/j.1749-6632.2011.06040.x

Geoffray, M.-M., Nicolas, A., Speranza, M., & Georgieff, N. (2016). Are circadian rhythms new pathways to understand Autism Spectrum Disorder? Journal of Physiology-Paris, 110(4), 434–438. https://doi.org/10.1016/j.jphysparis.2017.06.002

Goldman, S. E., Surdyka, K., Cuevas, R., Adkins, K., Wang, L., & Malow, B. A. (2009). Defining the Sleep Phenotype in Children With Autism. Developmental Neuropsychology, 34(5), 560–573. https://doi.org/10.1080/87565640903133509

Gotham, K., Pickles, A., & Lord, C. (2009). Standardizing ADOS Scores for a Measure of Severity in Autism Spectrum Disorders. Journal of Autism and Developmental Disorders, 39(5), 693–705. https://doi.org/10.1007/s10803-008-0674-3

Gottlieb, D. J., Somers, V. K., Punjabi, N. M., & Winkelman, J. W. (2017, March 1). Restless legs syndrome and cardiovascular disease: a research roadmap. Sleep Medicine, Vol. 31, pp. 10–17. Elsevier B.V. https://doi.org/10.1016/j.sleep.2016.08.008

Grandner, M., Mullington, J. M., Hashmi, S. D., Redeker, N. S., Watson, N. F., & Morgenthaler, T. I. (2018). Sleep duration and hypertension: Analysis of > 700,000 adults by age and sex. Journal of Clinical Sleep Medicine, 14(6), 1031–1039. https://doi.org/10.5664/jcsm.7176

Gringras, P., Gamble, C., Jones, A. P., Wiggs, L., Williamson, P. R., Sutcliffe, A., … Appleton, R. (2012). Melatonin for sleep problems in children with neurodevelopmental disorders: randomised double masked placebo controlled trial. BMJ, 345(nov05 1), e6664–e6664. https://doi.org/10.1136/bmj.e6664

Gringras, Paul, Nir, T., Breddy, J., Frydman-Marom, A., & Findling, R. L. (2017). Efficacy and Safety of Pediatric Prolonged-Release Melatonin for Insomnia in Children With Autism Spectrum Disorder. Journal of the American Academy of Child & Adolescent Psychiatry, 56(11), 948-957.e4. https://doi.org/10.1016/j.jaac.2017.09.414

Guénolé, F., Godbout, R., Nicolas, A., Franco, P., Claustrat, B., & Baleyte, J.-M. (2011). Melatonin for disordered sleep in individuals with autism spectrum disorders: Systematic review and discussion. Sleep Medicine Reviews, 15(6), 379–387. https://doi.org/10.1016/j.smrv.2011.02.001

Hollway, J. A., Aman, M. G., & Butter, E. (2013). Correlates and risk markers for sleep disturbance in participants of the autism treatment network. Journal of Autism and Developmental Disorders, 43(12), 2830–2843. https://doi.org/10.1007/s10803-013-1830-y

Léger, D., Guilleminault, C., Bader, G., Lévy, E., & Paillard, M. (2002). Medical and Socio-Professional Impact of Insomnia. Sleep, 25(6), 621–625. https://doi.org/10.1093/sleep/25.6.621

Lord, C. (2019). Taking Sleep Difficulties Seriously in Children With Neurodevelopmental Disorders and ASD. Pediatrics, 143(3). https://doi.org/10.1542/peds.2018-2629

Loy, J. H., Merry, S. N., Hetrick, S. E., & Stasiak, K. (2017). Atypical antipsychotics for disruptive behaviour disorders in children and youths. Cochrane Database of Systematic Reviews, 2017(8). https://doi.org/10.1002/14651858.CD008559.pub3

Manelis-Baram, L., Meiri, G., Ilan, M., Faroy, M., Michaelovski, A., Flusser, H., … Dinstein, I. (2021). Sleep Disturbances and Sensory Sensitivities Co-Vary in a Longitudinal Manner in Pre-School Children with Autism Spectrum Disorders. Journal of Autism and Developmental Disorders, (0123456789). https://doi.org/10.1007/s10803-021-04973-2

Maras, A., Schroder, C. M., Malow, B. A., Findling, R. L., Breddy, J., Nir, T., … Gringras, P. (2018). Long-Term Efficacy and Safety of Pediatric Prolonged-Release Melatonin for Insomnia in Children with Autism Spectrum Disorder. Journal of Child and Adolescent Psychopharmacology, 28(10), 699–710. https://doi.org/10.1089/cap.2018.0020

Mazurek, M. O., & Sohl, K. (2016). Sleep and Behavioral Problems in Children with Autism Spectrum Disorder. Journal of Autism and Developmental Disorders, 46(6), 1906–1915. https://doi.org/10.1007/s10803-016-2723-7

Meiri, G., Dinstein, I., Michaelowski, A., Flusser, H., Ilan, M., Faroy, M., … Menashe, I. (2017). Brief Report: The Negev Hospital-University-Based (HUB) Autism Database. Journal of Autism and Developmental Disorders, 47(9). https://doi.org/10.1007/s10803-017-3207-0

Miano, S., Bruni, O., Elia, M., Trovato, A., Smerieri, A., Verrillo, E., … Ferri, R. (2007). Sleep in children with autistic spectrum disorder: A questionnaire and polysomnographic study. Sleep Medicine, 9(1), 64–70. https://doi.org/10.1016/j.sleep.2007.01.014

Miller, B. J., & McCall, W. V. (2022). Insomnia and suicide as reported adverse effects of second-generation antipsychotics and mood stabilizers. Journal of Clinical Sleep Medicine, 18(2), 517–522. https://doi.org/10.5664/jcsm.9646

Nobre, B., Rocha, I., Morin, C. M., & Meira e Cruz, M. (2021). Insomnia and circadian misalignment: An underexplored interaction towards cardiometabolic risk. Sleep Science, Vol. 14, pp. 55–63. Brazilian Association of Sleep and Latin American Federation of Sleep Societies. https://doi.org/10.5935/1984-0063.20200025

Owens, J. A., & Maski, K. (2016). Insomnia, parasomnias, and narcolepsy in children: clinical features, diagnosis, and management. In Lancet Neurol (Vol. 15).

Reutrakul, S., & Van Cauter, E. (2018, July 1). Sleep influences on obesity, insulin resistance, and risk of type 2 diabetes. Metabolism: Clinical and Experimental, Vol. 84, pp. 56–66. W.B. Saunders. https://doi.org/10.1016/j.metabol.2018.02.010

Reynolds, A. M., Soke, G. N., Sabourin, K. R., Hepburn, S., Katz, T., Wiggins, L. D., … Levy, S. E. (2019). Sleep problems in 2-to 5-year-olds with autism spectrum disorder and other developmental delays. Pediatrics, 143(3). https://doi.org/10.1542/peds.2018-0492

Richdale, A. L., & Schreck, K. A. (2009). Sleep problems in autism spectrum disorders: Prevalence, nature, & possible biopsychosocial aetiologies. Sleep Medicine Reviews, 13(6), 403–411. https://doi.org/10.1016/j.smrv.2009.02.003

Rossignol, D. A., & Frye, R. E. (2011). Melatonin in autism spectrum disorders: a systematic review and meta-analysis. Developmental Medicine & Child Neurology, 53(9), 783–792. https://doi.org/10.1111/j.1469-8749.2011.03980.x

Samanta, P., Mishra, D. P., Panigrahi, A., Mishra, J., Senapati, L. K., & Ravan, J. R. (2020). Sleep disturbances and associated factors among 2-6-year-old male children with autism in Bhubaneswar, India. Sleep Medicine, 67, 77–82. https://doi.org/10.1016/j.sleep.2019.11.1244

Schroder, C. M., Banaschewski, T., Fuentes, J., Hill, C. M., Hvolby, A., Posserud, M.-B., & Bruni, O. (2021). Pediatric prolonged-release melatonin for insomnia in children and adolescents with autism spectrum disorders. Expert Opinion on Pharmacotherapy, 22(18), 2445–2454. https://doi.org/10.1080/14656566.2021.1959549

Schroder, C. M., Malow, B. A., Maras, A., Melmed, R. D., Findling, R. L., Breddy, J., … Gringras, P. (2019). Pediatric Prolonged-Release Melatonin for Sleep in Children with Autism Spectrum Disorder: Impact on Child Behavior and Caregiver’s Quality of Life. Journal of Autism and Developmental Disorders, 49(8), 3218–3230. https://doi.org/10.1007/s10803-019-04046-5

Sivertsen, B., Krokstad, S., Mykletun, A., & Øverland, S. (2009). Insomnia Symptoms and Use of Health Care Services and Medications: The HUNT-2 Study. Behavioral Sleep Medicine, 7(4), 210–222. https://doi.org/10.1080/15402000903190199

Skaer, T. L., & Sclar, D. A. (2010). Economic Implications of Sleep Disorders. PharmacoEconomics, 28(11), 1015–1023. https://doi.org/10.2165/11537390-000000000-00000

Sofi, F., Cesari, F., Casini, A., Macchi, C., Abbate, R., & Gensini, G. F. (2014). Insomnia and risk of cardiovascular disease: A meta-analysis. European Journal of Preventive Cardiology, 21(1), 57–64. https://doi.org/10.1177/2047487312460020

Sørensen, L., Jensen, M. S. A., Rathleff, M. S., & Holden, S. (2019). Comorbid insomnia, psychological symptoms and widespread pain among patients suffering from musculoskeletal pain in general practice: A cross-sectional study. BMJ Open, 9(6). https://doi.org/10.1136/bmjopen-2019-031971

Tzischinsky, O., Meiri, G., Manelis, L., Bar-Sinai, A., Flusser, H., Michaelovski, A., … Dinstein, I. (2018). Sleep disturbances are associated with specific sensory sensitivities in children with autism. Molecular Autism, 9(1), 1–10. https://doi.org/10.1186/s13229-018-0206-8

Vargas, I., Vgontzas, A. N., Abelson, J. L., Faghih, R. T., Morales, K. H., & Perlis, M. L. (2018). Altered ultradian cortisol rhythmicity as a potential neurobiologic substrate for chronic insomnia. Sleep Medicine Reviews, 41, 234–243. https://doi.org/10.1016/j.smrv.2018.03.003

Veatch, O. J., Sutcliffe, J. S., Warren, Z. E., Keenan, B. T., Potter, M. H., & Malow, B. A. (2017). Shorter sleep duration is associated with social impairment and comorbidities in ASD. Autism Research, 10(7), 1221–1238. https://doi.org/10.1002/aur.1765

Wechsler, D. (1967). Wechsler Preschool and Primary Scale of Intelligence. New York, NY: Psychological Corp.

