## Supplementary Tables for "Burden of insomnia on healthcare utilization in children with autism spectrum disorder"

**Table S1.** List of medication for the management of chronic conditions by main indication

|  | **Percent** |
| --- | --- |
| **Asthma/Pulmonary Disorders** | **15.80%** |
| Montelukast | 7.40% |
| Beclometasone | 1.30% |
| Formoterol and Beclometasone | 0.40% |
| Salbutamol | 0.40% |
| Symbicort/Duoresp | 0.20% |
| Fluticasone | 5.70% |
| Budesonide | 0.40% |
| **Attention Deficit Hyperactivity Disorder** | **13.40%** |
| Methylphenidate | 9.80% |
| amfetamine-Dexamfetamine | 1.70% |
| Lisdexamfetamine | 1.70% |
| Dexmethylphenidate | 0.20% |
| **Insomnia** | **11.80%** |
| Melatonin | 10.90% |
| Promethazine | 0.90% |
| **Mental and Mood Conditions** | **11.40%** |
| Risperidone | 3.90% |
| Aripiprazole | 2.60% |
| Periciazine | 2.20% |
| Clotiapine | 0.60% |
| Fluoxetine | 0.20% |
| Haloperidol | 0.20% |
| Quetiapine | 0.20% |
| Clonazepam | 1.30% |
| Biperiden | 0.20% |
| **Nutrient Deficiencies** | **6.90%** |
| Ferric Oxide Polymaltose complexes | 5.50% |
| Pyridoxine (Vitamin B6) | 0.60% |
| Cholecalciferol | 0.40% |
| Levocarnitine | 0.40% |
| **Food supplements** | **5.80%** |
| Enfamil AR 1 Lipil CD | 1.10% |
| Enfamil AR 2 Lipil CD | 0.90% |
| Nutren CD | 0.70% |
| Nutrilon AR | 0.70% |
| Nutramig_Lipil1/Nutrilon Pepti 1 | 0.60% |
| Nutramigen 2 CD | 0.40% |
| Nutrilon Pepti Junior | 0.40% |
| Neocate CD | 0.20% |
| Nutramig_Lipil2/Nutrilon Pepti 2 | 0.20% |
| Nutramigen 1 CD | 0.20% |
| Pediasure CD | 0.20% |
| Special Grass MIX | 0.20% |
| **Seizure Disorders** | **5.40%** |
| Valproic Acid | 1.80% |
| Levetiracetam | 1.70% |
| Phenobarbital | 0.90% |
| Cannabidiol | 0.40% |
| Gabapentin | 0.20% |
| Stiripentol | 0.20% |
| Topiramate | 0.20% |
| **Skin Conditions** | **2.50%** |
| Mometasone | 0.90% |
| Pimecrolimus | 0.60% |
| AQUEOUS CREAM CD | 0.20% |
| Clobetasone | 0.20% |
| Diflucortolone | 0.20% |
| Griseofulvin | 0.20% |
| EMOL/AQUAPHOR CD | 0.20% |
| **Allergic Disorders** | **1.80%** |
| Triamcinolone | 0.20% |
| Dimethindene | 0.60% |
| Desloratadine | 0.40% |
| Azelastine | 0.20% |
| Levocabastine | 0.20% |
| Epinephrine | 0.20% |
| **Endocrine and Metabolic Diseases** | **1.50%** |
| Levothyroxine Sodium | 0.70% |
| Glucagon | 0.20% |
| Glucose | 0.20% |
| Insulin Aspart | 0.20% |
| Insulin Degludec | 0.20% |
| **Gastrointestinal Disorders** | **1.40%** |
| Omeprazole | 0.40% |
| Ranitidine | 0.40% |
| Esomeprazole | 0.20% |
| Gaviscon Advance CD | 0.20% |
| Macrogol | 0.20% |
| **Pre-Surgical Medications** | **1.10%** |
| Midazolam | 0.90% |
| Atropine | 0.20% |
| **Infections** | **0.80%** |
| Dexamycin/Dethamycin CD | 0.20% |
| Ofloxacin | 0.20% |
| Tobramycin | 0.20% |
| Tarocidin/Phenimixin CD | 0.20% |
| **Urologic Disorders** | **0.60%** |
| Desmopressin | 0.40% |
| Oxybutynin | 0.20% |
| **Autoimmune diseases** | **0.40%** |
| Colchicine | 0.20% |
| Tacrolimus | 0.20% |
| **Cardiovascular Diseases** | **0.40%** |
| Propranolol | 0.20% |
| Spironolactone | 0.20% |
| **Ophthalmologic Conditions** | **0.40%** |
| Dexpanthenol-Sodium Hyaluronate CD | 0.20% |
| Pilocarpine | 0.20% |
| **Rheumatic diseases** | **0.20%** |
| Tetracosactide | 0.20% |
| Methotrexate | 0.20% |
| **Otorhinolaryngology Conditions** | **0.20%** |
| Clean-Ears CD | 0.20% |
| **Pain Medications** | **0.20%** |
| Oxycodone | 0.20% |
| **Various Main Indications** | **0.20%** |
| Acetazolamide | 0.20% |
